# Are abnormalities in lipid metabolism, together with adverse childhood experiences, the silent causes of immune-linked neurotoxicity in major depression?

**DOI:** 10.1101/2024.02.20.24302841

**Authors:** Michael Maes, Ketsupar Jirakran, Asara Vasupanrajit, Bo Zhou, Chavit Tunvirachaisakul, Drozdstoj St. Stoyanov, Abbas F. Almulla

## Abstract

**Background:** Severe or recurring major depression is associated with increased adverse childhood experiences (ACEs), heightened atherogenicity, and immune-linked neurotoxicity (INT). Nevertheless, the interconnections among these variables in outpatient of major depression (OMDD) have yet to be determined.

**Objectives:** Determine the correlations among INT, atherogenicity, and ACEs in 66 OMDD patients (of whom thirty-three had metabolic syndrome, MetS) and sixty-seven controls (31 of whom had MetS).

**Results:** The free cholesterol/reverse cholesterol transport ratio, apolipoprotein (Apo) B and E, and a comprehensive atherogenicity index were all significantly associated with increased INT in OMDD subjects without MetS. ACEs were substantially correlated with INT in patients with MetS. INT (only in MetS) and atherogenicity indices (only in people without MetS) were significantly associated with the clinical phenome features of OMDD, including the recurrence of illness (ROI, including lifetime suicidal behaviors), the lifetime phenome (neuroticism + lifetime anxiety disorders and dysthymia), and the current phenome (including current suicidal behaviors). A significant proportion of the variability (58.3%) in the lifetime + current phenome could be accounted for by INT, interactions between INT and atherogenicity (labeled "atherommune index"), ApoE, three ACE subtypes (all positively correlated), and age (inversely correlated). A single validated latent construct could be extracted from ROI, lifetime phenome, current phenome, INT, and atherommune index. 36.1% of this factor’s variance was accounted for by three ACE subtypes.

**Discussion:** We have developed a novel OMDD model, namely a pathway phenotype, labeled the "atherommune-phenome," which demonstrates that the interplay between INT and atherogenicity is essential to OMDD.

## Introduction

It is believed that major depressive disorder (MDD) is a neuroimmune disorder accompanied with dysfunctions in metabolic pathways. MDD is distinguished by a variety of immune disturbances, the specific manifestations of which may vary. It is observed that inpatients diagnosed with MDD (IMDD), the majority of whom suffer from very severe MDD and melancholia, exhibit indications of an immune response, including activation of M1 macrophages, T helper (Th)1, Th2 and Th17 immune cells, and an acute phase response with increased levels of acute phase proteins, complement factors, and pro-inflammatory cytokines, e.g., interleukin (IL)-6, IL-1β, and tumor necrosis factor (TNF)-α [1–3]. These findings have been extensively replicated and published in various meta-analyses [4–8].

In addition, severe first-episode (FE)-IMDD is characterized by elevated levels of IL-16, Th1 polarization, various chemokines (CCl2, CCL4, CCL11, CXCL8) and growth factors, such as platelet derived growth factor (PDGF) [9]. It is worth noting that severe IMDD is additionally distinguished by heightened concentrations of negative immunoregulatory and anti-inflammatory cytokines and their receptors, such as IL-10, IL-4, sIL-2 receptor (sIL-2R), and sIL-1R antagonist (sIL-1RA) [9]. These cytokines and their receptors are components of a broader compensatory immunoregulatory system (CIRS), which functions to dampen activated immune response system (IRS) activity thereby preventing hyperinflammation [10, 3].

Furthermore, it is now apparent that IMDD is distinguished by aberrations in lipid metabolism, which include a) reduced lecithin-cholesterol acyltransferase (LCAT), paraoxonase 1 (PON1) activity, and high-density lipoprotein (HDL) cholesterol levels, which lead to decreased reverse cholesterol transport (RCT), and b) increased atherogenicity, as measured by the atherogenicity index of plasma (AIP), Castelli risk index 1 and 2, ApoB/ApoA ratio, and newly developed indices [11–14]. These findings were recently confirmed in a meta-analysis that included 176 studies, involving 17,094 patients with affective disorders and 16,957 healthy controls [15].

Significantly, the impact of adverse childhood experiences (ACEs) on the “phenome” of severe IMDD is partially mediated by neuro-immune pathways, such as the activation of Th-1 and M1 immune cells [16]. The phenome of depression is a complex condition that encompasses the intensity of current depressive symptoms, anxiety, and suicidal tendencies [17, 18, 14]. This implies that the activation of the neuro-immune system by ACEs has an impact on the phenome of depression [15]. As described previously, these effects can be attributed to the heightened immune-linked neurotoxicity (INT) caused by M1, Th1, and Th17 cytokines and chemokines that may impact astroglial and neuronal projections, neurogenesis, and neuroplasticity [19–22].

However, different phenotypes of MDD may exhibit distinct neuroimmune disorders. In a study conducted by Maes et al. [23], it was found that young outpatients with very mild FE-MDD (FE-OMDD) have notably reduced levels of CIRS products including IL-4, IL-10, sIL-2R, and IL-12p40, whereas no activation of M1, Th1 or Th2 lineages could be detected. A pre-proatherogenic state was identified in the latter group of patients, despite the absence of subclinical metabolic syndrome (MetS) symptoms [24]. Young FE-OMDD patients exhibited elevated levels of free cholesterol and decreased LCAT activity, thus pointing towards a pre-proatherogenic state even in the absence of any signs of MetS [24] .

The neuroimmune profile of a more representative sample of OMDD patients with a broader spectrum of recurrent depression is once more distinct. These OMDD patients exhibit two distinct groups of proteins that are expressed differently. The first group consists of upregulated proteins, namely *CCL11, TNFB, PDGF, CCL9, IL4, CCL5, CCL2, CCL4*, and *IL1RN.* These proteins are all part of the same latent construct and form an intricately connected protein-protein interaction (PPI) network [25]. The second group consists of downregulated proteins, including *VEGFA, CCL3, CSF1, IL10*, and NGF [25]. Gene ontology (GO) annotation and enrichment analysis reveals that the subset of upregulated differentially expressed proteins (DEPs) is associated with GO functions including immune-inflammatory, defense, or stress responses. On the other hand, the second set of DEPs is linked to the regulation of neuronal death and neurogenesis [25]. Consequently, OMDD is characterized by changes in immune networks that may lead to heightened INT [25]. Furthermore, it is crucial to note that, in these OMDD patients, substantial interactions between OMDD and MetS significantly elevate immune-associated neurotoxicity, including TNF signaling, *IL17, CCL2*, and *CCL5*, and growth factors, such as *PDGF*, that potentially stimulate the upregulated DEP network [25].

In OMDD patients, a significant portion of the variation seen in suicidal behaviors (23.6%), neuroticism (26.6%), and the severity of the OMDD phenome (31.4%) was accurately predicted by INT and interaction between MetS and specific cytokines/chemokines/growth factors, such as CCL5, TNFB, or VEGFA. Maes et al. [25] observed increases in atherogenicity indices and decreases in RCT, LCAT, and HDL cholesterol in the same OMDD patients. Once more, increased atherogenicity indices partially mediated the effects of ACEs on the phenome of depression [14]. Two potential explanations exist for the disparities between FE-OMDD and the more representative sample of OMDD patients. First, it is crucial to consider the disparity in the recurrence of illness (ROI) index between the representative OMDD cohort, which comprises patients with a median of 3.9 episodes (range: 1-20) versus FE-OMDD. ROI is strongly associated with numerous biomarkers, including cytokine/chemokine/growth factor production [21, 18] and increased atherogenicity [16], and predicts the severity of the current phenome, including suicidal behaviors and neuroticism [26, 27]. Young FE-OMDD patients were, secondly, recruited at a younger age than individuals comprising the representative OMDD sample.

Despite this, information regarding the relationship between ACEs, increased atherogenicity, and INT is currently unavailable. Numerous studies have demonstrated that lipid metabolism influences and regulates immunity. An illustration of this can be seen in the potential influence of cholesterol homeostasis on innate and adaptive immune cells, autoimmunity, and anti-inflammatory response regulation [28–31]. Additionally, hypercholesterolemia has been associated with inflammatory responses [32], and immune functions are influenced by ApoE concentrations [33]. Furthermore, information regarding the potential effects of cumulative impacts of ACEs, INT, and modified lipid metabolism on the OMDD phenome, which includes ROI, neuroticism, and suicidal behaviors, is currently unavailable.

Hence, the current study was conducted to investigate the associations between ACEs, lipid concentrations and derived atherogenicity indices and whether those associations are differently regulated in participants with and without MetS. Secondly, we aim to examine whether immune-associated neurotoxicity combined with increased atherogenicity and ACEs is associated with the phenome of OMDD, comprising lifetime and current suicidal behaviors, ROI (based on number of episodes and suicidal behaviors), the lifetime phenome features (based on the presence of dysthymia, anxiety disorders and neuroticism), and the severity of the current phenome (based on depression and anxiety ratings and current suicidal behaviors).

## Methods and Participants

### Participants

This study comprised sixty-seven healthy controls and 66 OMDD patients. The control group was recruited during the period from September 2021 to February 2022 via oral communication within the catchment area, specifically in Bangkok, Thailand. Volunteers in good health comprised personnel, relatives, staff colleagues, and acquaintances of MDD patients. OMDD patients were enlisted as outpatients of the Department of Psychiatry at King Chulalongkorn Memorial Hospital in Bangkok, Thailand. The outpatients were diagnosed with MDD using the criteria outlined in the DSM-5 [34]. Our OMDD sample is representative of the Bangkok OMDD population due to the following: a broad recurrence of illness (ROI) index ranging from 1 to 20 episodes; a broad Hamilton Depression Rating Scale score range of 7 to 33 (including non-responders and partial remitters to treatment); a female to male sex ratio of 2.66; and a mean age of 36.9 (SD=11.5) years. In order to examine the impact of MetS on immune profiles, we recruited around 50% of the controls and OMDD samples to have a diagnosis of MetS. As such, sixty-four participants were diagnosed with MetS, while sixty-nine did not have the condition. Of the sixty-seven controls, thirty-three were diagnosed with MetS, and of the 66 OMDD patients, thirty-one were diagnosed with MetS.

The following are the exclusion criteria applicable to both patients and controls: a) psychiatric disorders other than MDD and classified as axis-1, encompassing substance use disorders (SUD) (excluding tobacco use disorder), schizophrenia, schizo-affective psychosis, bipolar disorder, autism spectrum disorders, and psycho-organic syndromes; b) disorders falling under axis-2, including borderline personality disorder and antisocial personality disorder; c) pregnant or lactating women, d) neurological disorders including stroke, Alzheimer’s and Parkinson’s disease, epilepsy, multiple sclerosis, and brain tumors. In addition, individuals who met the inclusion criteria for the control group and had a positive familial history of mood disorders, suicide, substance use disorders, or suffered from current and/or lifetime MDD and dysthymia were excluded from the research.

All participants voluntarily provided written informed consent prior to their involvement in the study. The research was conducted in compliance with ethical standards both internationally and in Thailand, as well as privacy laws. The inquiry was granted authorization (#445/63) by the Institutional Review Board of the Faculty of Medicine at Chulalongkorn University located in Bangkok, Thailand.

### Clinical assessments

Through the administration of a semi-structured questionnaire and interviews with patients and controls, sociodemographic and clinical data were gathered. The semi-structured interview comprised the following demographic questions: age, sex, years of education, marital status, employment status, occupation, income, number of depressive episodes, and familial anamnesis in first-degree family members. Furthermore, the interview investigates the applicant’s medical history and utilization of psychotropic and medical medications. MDD was diagnosed using the Mini International Neuropsychiatric Interview (M.I.N.I.), translated into Thai by Khamwongpin and Kittirattanapaiboon [35], and the DSM-5 criteria [35, 34]. The MINI was also used to make the diagnoses of other axis-1 diagnoses and to exclude patients and controls [35]. The severity of MDD was evaluated using the Beck Depression Inventory II (BDI-II) [36] and the Hamilton Rating Scale for Depression (HAMD) [37]. The BDI-II was translated into Thai [38]. A BDI subscore for “pure depression” was computed by summing the scores of the items that assess the cognitive aspects of depression. As a consequence, the items utilized to assess sex drive, appetite, weight loss, fatigue, irritability, and insomnia were excluded from the subscore sum. Five personality traits were assessed using a Thai translation of the Big Five Inventory (BFI) [39, 40].

In the present study, the neuroticism scores were employed, as they were found to be a substantial factor in the phenome of MDD[27]. Utilizing the Columbia Suicide Severity Rating Scale (C-SSRS) [41], the severity of previous and current suicidal thoughts and attempts was assessed. Utilizing the C-SSRS, the frequency, severity, intensity, and lethality of suicidal ideation (SI) and attempts (SA) were evaluated. “Current” (Current_SI or Current_SA) was considered as the symptoms present during last month before admission into the study, and lifetime (LT_SI and LT_SA) as the symptoms that occurred earlier. The assessment of ACEs was conducted utilizing a Thai translation of the ACE Questionnaire [42]. The survey comprises twenty-eight items that pertain to traumatic experiences during childhood and spans ten domains. For the purposes of this research, the raw summed scores on the following were utilized: emotional abuse, physical abuse, sexual abuse, domestic violence, substance abuse within the household, mental illness within the household, parental divorce, and a criminal household member. The present investigation also incorporated the total number of ACEs.

The indices that represent the OMDD phenome (ROI, suicidal behaviors, severity of the phenome) were computed based on the rating scale assessments. a) The Current_SI and LT_SI scores were computed using the first principal component extracted from seven items that assessed suicidal ideation on the C-SSRS; the Current_SA and LT_SA scores were computed as the first principal component extracted from five items that evaluated suicidal attempts on the C-SSRS [14]. Current_SB was subsequently represented by the sum of the z-transformations of the Current_SI and Current_SA scores, whilst LT_SB was the sum of the z scores of LT_SI and LT_SA. The composite score of the 4 different SB ratings was considered to reflect overall SB (labeled as Current+LT_SB). b) In order to compute the ROI index, the number of depressive episodes, the LT_SB and LT_SI scores were utilized to derive the first principal component. c) The severity of the current OMDD phenome was defined [14] as the first PC derived from the pure BDI score, total HAMD, and Current_SB score. d) the severity of the lifetime phenome was constructed as a principal component extracted from the neuroticism score, the presence of lifetime dysthymia, and the number of lifetime anxiety disorders [14]. As explained in the statistics section, prespecified quality criteria were rigorously met by each of these PCs.

When diagnosing TUD, the diagnostic criteria outlined in the DSM-5 were utilized. The body mass index (BMI) was calculated using the formula weight (kg) divided by height (m) squared. MetS was defined in accordance with the 2009 Joint Scientific Statement of the American Heart Association/National Heart, Lung, and Blood Institute [43] as the presence of three or more of the following components: a) a low HDL cholesterol level of less than 40 mg/dL for men and less than 50 mg/dL for women; b) a triglyceride level of 150 mg/dL; c) a waist circumference of at least 90 cm for men and 80 cm for women, or a BMI exceeding 25 kg/m^2^; d) an increase in fasting glucose level to 100 mg/dL or a diagnosis of diabetes; and e) a blood pressure exceeding 130 mm Hg systolic blood pressure or 85 mm Hg diastolic blood pressure.

### Analyses or assays

Between 8:00 and 9:00 a.m., we obtained twenty-five milliliters of fasting venous blood from each participant in the study using a serum tube and disposable syringe. Following, the blood underwent centrifugation at 3500 revolutions per minute, the serum was obtained and stored in Eppendorf containers containing aliquots at -80 degrees Celsius until it was defrosted in preparation for biomarker assays. Forty-eight cytokines, chemokines, and growth factors were analyzed for their fluorescence intensities (FI) utilizing the Bio-Plex Multiplex Immunoassay reagent supplied by Bio-Rad Laboratories Inc., Hercules, USA. The electronic supplementary file (ESF) comprises a comprehensive inventory of the analytes employed in our study, with their respective gene IDs and aliases listed in Table 1. Our methodology consisted of the following components: In order to dilute the serum to a ratio of 1:4, sample diluent (HB) was utilized. b) Following dilution, the specimens were transferred to a 96-well plate where 50µl of a microparticle cocktail (including cytokines, chemokines, and growth factors) was contained in each well. Following that, the plate was incubated at room temperature for one hour while being agitated at 850 rpm. c) Following three washes, 50µl of diluted Streptavidin-PE was added to each well of the plate, and the mixture was agitated at room temperature for ten minutes. d) The subsequent analysis for assessing cytokines, chemokines, and growth factors was conducted utilizing the Bio-Plex® 200 System manufactured by Bio-Rad Laboratories, Inc., subsequent to the transfer of the sample to the 96-well plate. It was determined that the intra-assay CV values for each analyte were all below 11.0%. The standard concentrations supplied by the manufacturer were utilized to determine the concentrations of the samples. Subsequently, the proportion of concentrations falling below the minimum measurable concentration (referred to as "out of range" or "OOR") was computed. In ESF, Table 1, the quantity of analytes that demonstrated quantifiable values (i.e., those exceeding OOR) is specified in detail. In order to avoid analytes with OORs exceeding 60%, IL-2, IL-3, IL-5, IL-6, IL-7, and IL-12p70 were excluded. Analytes exhibiting quantifiable concentrations within the range of 40% to 60% were considered as prevalences and were transformed into dummy variables. As a result, IL-10 was dichotomized (measurable versus not) and utilized in place of the actual FI values. The other analytes were utilized as blank-subtracted FI values rather than the absolute concentrations in our statistical analysis due to their greater accuracy, particularly when multiple plates are employed. Previously, we extracted different principal components from the differently expressed proteins (DEPs), namely from the upregulated DEPs we extracted PC05 and PC06, and from the downregulated DEPs we extracted another PC.

**Table 1.** Socio-demographic, clinical, and biomarker data in outpatients with a major depressive episode (OMDD) and healthy controls (HC). All data are shown as mean (SD) or as ratio. All results of analysis of variance (F values), analysis of contingency analysis (Χ^2^ test), or MWU: Mann-Whitney U test. BMI: body mass index, MetS: metabolic syndrome, HAM-D: Hamilton Depression Rating Scale score, BDI: Beck Depression Inventory, ROI: recurrence of illness index, SB: suicidal behaviors, LT_ lifetime, PC05 and PC06: principal components (PC) extracted from 20 and 10 cytokines/chemokines/growth factors, respectively. PCdown: a PC extracted from five downregulated proteins, INT: immune-linked neurotoxicity, PRO/ANTI: a composite score reflecting a comprehensive atherogenicity index, FC/RCT: a composite score reflecting the ratio of free cholesterol on reverse cholesterol transport, atherommune: a composite score based on INT and PRO/ANTI measurements.

| Variables | HC (n=67) | OMDD (n=66) | F/X2 | df | p |
| --- | --- | --- | --- | --- | --- |
| Age (years) | 37.9 (9.2) | 37.0 (11.5) | 0.26 | 1/131 | 0.609 |
| Sex (F/M) | 58/9 | 48/18 | 3.94 | 1 | 0.055 |
| Education (years) | 14.1 (3.4) | 14.8 (2.8) | 1.72 | 1/131 | 0.193 |
| Number of episodes | - | 3.9 (range: 1-20) | - | - | - |
| BMI (kg/m2) | 27.4 (6.1) | 26.9 (5.4) | 0.27 | 1/130 | 0.607 |
| MetS (No/Yes) | 34/33 | 35/31 | 0.07 | 1 | 0.792 |
| TUD (No/Yes) | 62/5 | 54/12 | 3.43 | 1 | 0.064 |
| Total HAMD score | 2.3 (3.0) | 18.1 (6.4) | 330.94 | 1/131 | <0.001 |
| Total BDI-II score | 5.6 (6.5) | 26.9 (13.8) | 129.52 | 1/131 | <0.001 |
| Pure BDI (z score) | -0.687 (0.376) | 0.697 (0.952) | 122.20 | 1/131 | <0.001 |
| Total number of ACEs | 1.1 (1.5) | 3.0 (2.3) | MWU | - | <0.001 |
| ROI (z score) | -0.752 (0.0) | 0.776 (0.918) | MWU | - | <0.001 |
| Neuroticism | 18.3 (5.0) | 27.2 (6.4) | 80.33 | 1/131 | <0.001 |
| LT_phenome (z score) | -0.596 (0.299) | 0.605 (1.096) | MWU | - | <0.001 |
| LT_SB (z scores) | -0.554 (0.0) | 1.181 (0.146) | MWU | - | <0.001 |
| Current Suicidal behaviors (SI and SA) | -0.420 (0.0) | 0.426 (1.290) | MWU | - | <0.001 |
| Current_Phenome (z scores) | -0.749 (0.422) | 0.760 (0.828 ) | MWU | - | <0.001 |
| LT+Current_SB (z scores) | -0.534 (0.0) | 0.550 (1.200) | MWU | - | <0.001 |
| LT+Current_Phenome (z scores) | -0.719 (0.233) | 0.741 (0.944) | MWU | - | <0.001 |
| PC05 (z scores) | -0.291 (0.944) | 0.296 (0.970) | 12.48 | 1/131 | 0.001 |
| PC06 (z scores) | -0.318 (0.924) | 0.323 (0.969) | 13.65 | 1/131 | <0.001 |
| PCdown (z scores) | 0.320 (0.947) | -0.324 (0.945) | 15.42 | 1/131 | <0.001 |
| INT index (z score) | -0.488 (0.744) | 0.496 (0.978) | 42.71 | 1/131 | <0.001 |
| PRO/ANTI (z score) | -0.099 (1.167) | 0.101 (0.792) | 0.34 | 1/131 | 0.249 |
| PRO/ANTI (z scores) No MetS | -0.392 (0.163) | 0.384 (0.163) | 11.00 | 1/62 | 0.002 |
| FC/RCT (z scores) | -0.085 (1.079) | 0.086 (0.905) | 0.984 | 1/131 | 0.323 |
| FC/RCT (z scores) No MetS | -0.301 (0.950) | 0.292 (0.973) | 6.57 | 1/131 | 0.013 |
| ApoE | 977.3 (457.2) | 1096.6 (430.4) | 2.40 | 1/131 | 0.124 |
| Atherommune | -0.574 (1.337) | 0.582 (1.332) | 24.93 | 1/131 | <0.001 |

PC05 loaded highly (loadings >0.5) on 20 cytokines, chemokines, and growth factors, which were shown (using PPI analysis) to shape a tight PPI network [25]. Moreover, annotation analysis showed that their PPI network was enriched in the following GO functions: immune and inflammatory response, cytokine-mediated signaling pathway, defense response, and chemotaxis. This PC05 network included: G-CSF, CXCL1, HGF, sIL-1RA, sIL-2R, IL-4, IL-8, IL-9, IL-16, IL-17, CCL2, MIF, CCL3, CCL4, PDGF, CCL5, SCGF, CXCL12, TNF-α, and TNF-β. PC06 loaded highly (>0.6) on 10 cytokines, chemokines and growth factors and PPI analysis showed that these 10 immune variables shaped a tight PPI network [25], comprising sIL-1RA, IL-4, IL-9, IL-17, CCL4, PDGF, CCL5, CXCL12, TNF-α, and TNF-β. Moreover, annotation analysis showed that their PPI network was enriched in the following GO functions: chemotaxis, defense response, immune response, and response to stress [25]. It should be added that PC06 complied with all prespecified criteria of a validated latent structure, indicating that the ten immune compounds of PC06 are manifestations of a tight PPI network that causes specific dysfunctions. From five downregulated DEPs (namely: CCL3, CSF1, VEGF, NGF, and IL-10) we extracted another validated principal component (labeled PCdown), and annotation analysis showed that the PPI network of these downregulated DEP network was enriched in regulation of neuron death, regulation of microglial cell migration, positive regulation of neurogenesis. Based on the PC scores of PC06 and PC neurogenesis we built a new z unit-based composite, namely PC06 – PCdown, which reflects immune-linked neurotoxicity (labeled as INT).

As previously described [14, 24], the concentrations of total cholesterol, HDL-cholesterol, triglycerides, and direct LDL-cholesterol were ascertained using the Alinity C (Abbott Laboratories, USA; Otawara-Shi, Tochigi-Ken, Japan). The coefficients of variation for triglycerides, HDL-cholesterol, LDL-cholesterol, and total cholesterol were 4.5%, 2.6%, 2.3%, and 2.3%, respectively. The determination of ApoA1 and ApoB concentrations was performed using immunoturbidimetric assays that involved the Roche Cobas 6000 and c501 modules (Roche, Rotkreuz, Switzerland). Apo A1 and Apo B exhibited intra-assay coefficients of variation of 1.75 and 2.64 percent, respectively. Using the Free Cholesterol Colorimetric Assay reagent (Elabscience, cat number: E-BC-K004-M) and the Human ApoE (Apolipoprotein E) ELISA reagent, the amounts of free cholesterol and ApoE were determined, respectively. The respective intra-assay coefficients of variation were 1.9% and 4.67%, respectively. The esterified cholesterol ratio, which serves as a measure of LCAT activity, was computed using the formula (1-free cholesterol / total cholesterol) x 100 (1). The ratio of ApoB to ApoA was computed in order to quantify atherogenicity. Additional atherogenicity indices were computed by Maes et al. [14] as a z-unit-based composite score: z transformation of ApoB (z ApoB) + z triglycerides + z free cholesterol + z LDL-cholesterol (labeled PRO). Moreover, an RCT index (designated RCT) was computed as follows: z HDL-cholesterol + z LCAT + z ApoA1. Two atherogenicity indices were calculated: a) the PROATH/RCT ratio, which was defined as z triglycerides + z ApoB + z free cholesterol + z LDLc - z HDLc - z LCAT - z ApoA1; and b) the free cholesterol /RCT composite ratio, computed as z free cholesterol – z RCT. Since these ratios do not contain ApoE, we used three lipid variables in our analysis, namely both abovementioned ratios and ApoE.

### Statistical analysis

In order to determine the correlations between two sets of scale variables, Pearson’s product moment or Spearman’s rank order coefficients were applied. To analyze scale variables among diverse diagnostic groups, analysis of variance was employed. In order to compare nominal variables across distinct categories, the chi-square test or Fisher’s Exact Probability Test was utilized. No false discovery rate (FDR) p-correction was implemented on the immune or lipid data during analysis, including for multiple comparisons or correlations. In fact, statistical analyses cannot regard these variables to be independent due to their intricate networks of interactions. An investigation was conducted into the relationships between explanatory variables (ACEs, immune and lipid data) and dependent variables (clinical phenome data) using multiple regression analysis. We used a manual method and additionally, in order to ascertain the most precise predictors for the model, we utilized a forward stepwise automatic regression method employing p-values of 0.05 for inclusion and 0.06 for exclusion. Standardized coefficients, t-statistics, and exact p-values were included in the final regression models for each of the explanatory variables, along with total variance (represented by R^2^ or partial eta squared as effect size) and F statistics (and p values). Collinearity and multicollinearity were analyzed utilizing tolerance (cut-off value <0.25), the variance inflation factor (cut-off value >4), condition index, and variance proportions from the collinearity diagnostics table. In order to determine whether heteroskedasticity was present, the White and modified Breusch-Pagan test was utilized.

In order to reduce the number of variables to a singular PC score (feature reduction) that could be utilized in subsequent statistical analyses, PC analysis (PCA) was performed. To determine factorability, the Bartlett’s sphericity test and the Kaiser-Meyer-Olkin test for sample adequacy were utilized. In order to develop clinical PC models that are reflective of a latent construct, the first PC extracted from all variables was considered acceptable only if the variance explained (VE) surpassed 50%, all loadings on the initial PC surpassed 0.6, and Cronbach’s alpha surpassed 0.7. The data distributions underwent the necessary normalization processes, which included rank-based, logarithmic, square-root, and inverse normal transformations. In addition, z-score transformation was applied to the data in order to improve their interpretability and produce z-unit-weighted composite scores that represent distinct immune profiles more precisely. A two-tailed design was employed in each of the aforementioned investigations, with statistical significance being determined at an α value of 0.05. The software utilized was IBM Windows SPSS version 29.

The a priori sample size was computed utilizing G*Power 3.1.9.4. The principal statistical analysis that was pre-established was a multiple regression analysis, with the LT+Current phenome score as dependent variable and immune and lipid data serving as input variables. Given f = 0.136 (which accounts for approximately 12% of the explained variance), 5 explanatory variables, an alpha value of 0.05, and a power of 0.8, the bare minimum sample size was 100.

## Results

### Sociodemographic, clinical and biomarker data in OMDD and controls

**Table 1** shows the sociodemographic, clinical, and biomarker data in OMDD patients versus healthy controls. There were no significant differences in age, sex, education, BMI, MetS and TUD between the study samples. All clinical scores were significantly higher in OMDD patients than in controls. PC05 and PC06 scores were significantly higher in OMDD than in controls, whereas PCdown was significantly lower on OMDD than controls. The INT index was significantly higher in OMDD than controls. There was no difference in the PRO/ANTI ratio among OMDD and controls. Nevertheless, this ratio was increased in OMDD patients without MetS as compared with controls without MetS.

### Association of neuroimmune networks and lipid data

**Table 2** shows the results of multiple regression analyses with the neuroimmune PCs as dependent variables and lipid data, BMI, and confounding variables as explanatory variables. The regressions were repeated in subjects without MetS (the “a” labeled regressions) and in subjects with MetS (“b” labeled regressions). Regressions #1a shows that, in subjects without MetS, 22.2% of the variance in PC05 was explained by the regression on FC/RCT and BMI (both positively associated). On the other hand, in subjects with MetS, 25.6% of the variance in PC05 was explained by the regression on emotional and sexual abuse. Regression 2a shows that the cumulative effects of FC/RCT and BMI explained 20.3% of the variance in PC06 in individuals without MetS. In those with MetS (regression 2b) 26.6% of the variance in PC06 was explained by the two ACE subtypes as described above. We found that 7.8% of the variance in PCdown in subjects without MetS was explained by ACE neglect (inversely associated). 19.6% of the variance in the INT index in people without MetS was explained by BMI, ApoB and emotional neglect (all positively associated). 16.6% of the variance in the INT index in those with MetS was explained by emotional neglect and sexual abuse.

**Table 2.** Results of multiple regression analysis with immune network data as dependent variables. PC05 and PC06: principal components (PC) extracted from 20 and 10 cytokines/chemokines/growth factors, respectively. PCdown: a PC extracted from five downregulated proteins, INT: immune-linked neurotoxicity, FC/RCT: a composite score reflecting the ratio of free cholesterol on reverse cholesterol transport, MetS: metabolic syndrome, Em: emotional, Sex: sexual.

| Dependent Variables | Explanatory variables | B | T | p | R<sup>2</sup> | F | df | P |
| --- | --- | --- | --- | --- | --- | --- | --- | --- |
| 1a. PC05 NoMetS | Model |  |  |  | 0.222 | 9.26 | 2/65 | <0.001 |
|  | FC/RCT | 0.325 | 2.95 | 0.004 |  |  |  |  |
|  | BMI | 0.301 | 2.73 | 0.008 |  |  |  |  |
| 1b. PC05 MetS | Model |  |  |  | 0.256 | 10.31 | 2/60 | <0.001 |
|  | EmAbuse | 0.404 | 3.61 | 0.001 |  |  |  |  |
|  | SexAbuse | 0.274 | 2.45 | 0.017 |  |  |  |  |
| 2a. PC06 NoMetS | Model |  |  |  | 0.203 | 8.26 | 2/65 | 0.001 |
|  | FC/RCT | 0.339 | 3.03 | 0.003 |  |  |  |  |
|  | BMI | 0.257 | 2.30 | 0.025 |  |  |  |  |
| 2b. PC06 MetS | Model |  |  |  | 0.266 | 10.89 | 2/60 | <0.001 |
|  | EmAbuse | 0.420 | 3.79 | <0.001 |  |  |  |  |
|  | SexAbuse | 0.267 | 2.41 | 0.019 |  |  |  |  |
| 3a. PCdown NoMetS | Model |  |  |  | 0.078 | 5.67 | 1/66 | 0.020 |
|  | EmNeglect | -0.279 | -2.38 | 0.020 |  |  |  |  |
| 4a. INT NoMetS | Model |  |  |  | 0.196 | 5.20 | 3/64 | 0.003 |
|  | BMI | 0.283 | 2.53 | 0.014 |  |  |  |  |
|  | ApoB | 0.238 | 2.12 | 0.038 |  |  |  |  |
|  | EmNeglect | 0.227 | 2.03 | 0.047 |  |  |  |  |
| 4b. INT MetS | Model |  |  |  | 0.166 | 5.96 | 2/60 | 0.004 |
|  | EmNeglect | 0.302 | 2.56 | 0.013 |  |  |  |  |
|  | SexAbuse | 0.258 | 2.19 | 0.033 |  |  |  |  |

### Associations between immune and atherogenicity and clinical data

**Table 3** shows the correlations (Pearson correlation coefficients) between immune (PC06 and INT) and atherogenicity (PRO/ANTI) and clinical data. In those without MetS, PC06 was significantly correlated with the PRO/ANTI ratio but not with clinical data. In those with MetS, PC06 and INT were significantly correlated with all clinical data, but not with the PRO/ANTI ratio. Moreover, the INT index was associated with ROI, neuroticism, and the phenome. The PRO/ANTI ratio was significantly associated with all clinical data (except BDI) but only in subjects without MetS. The total ACE score was correlated with all clinical data (except Current_SB in those with MetS) in both people with and without MetS.

**Table 3.**
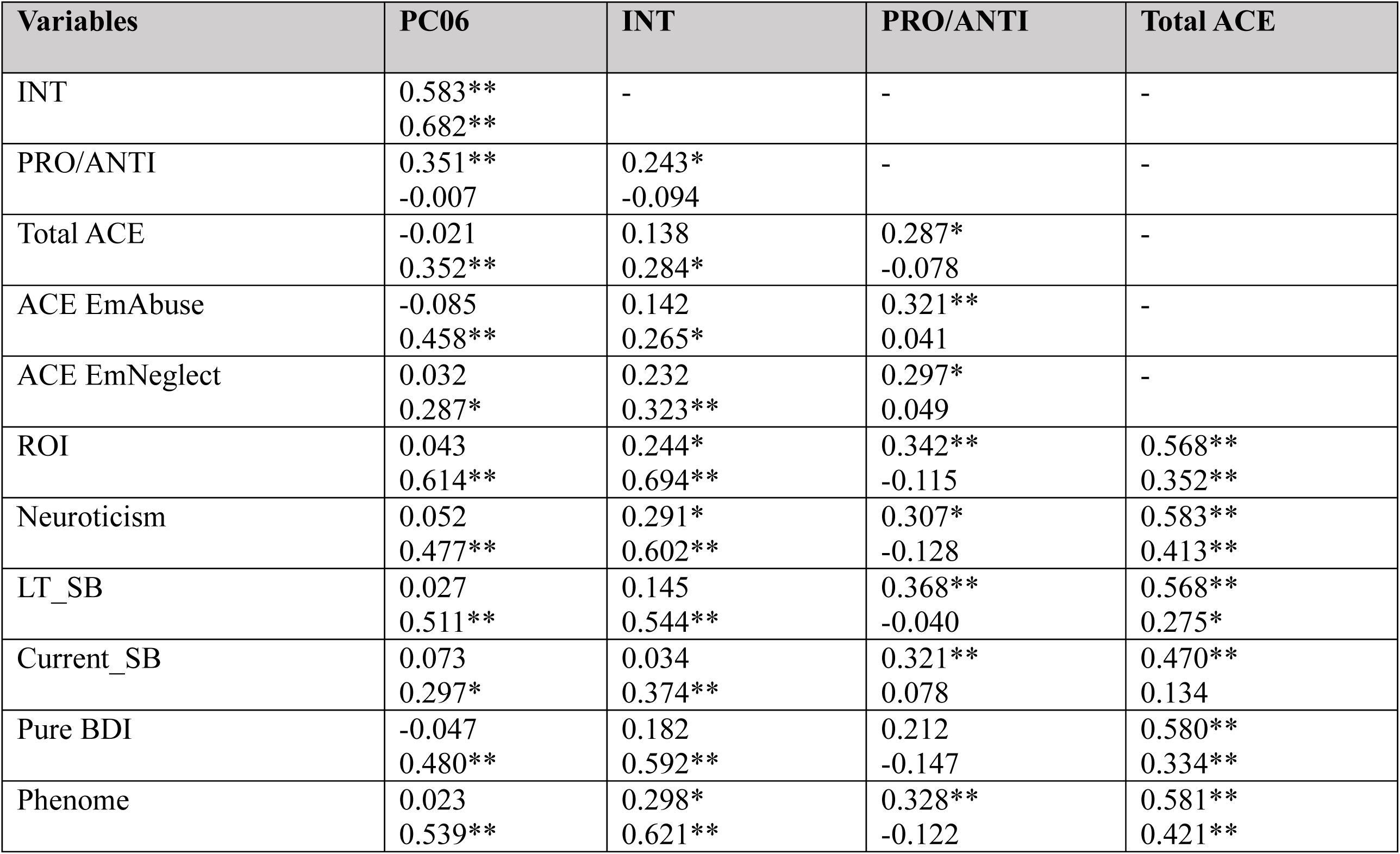
Correlation matrix among immune, atherogenicity and clinical data * p<0.05, ** p<0.01 (two tailed). Upper figures: in the subgroup without metabolic syndrome; lower figures: in the group with metabolic syndrome. PC06: principal components (PC) extracted from 10 cytokines/chemokines/growth factors. INT: immune-linked neurotoxicity, PRO/ANTI: a composite score reflecting a comprehensive atherogenicity index, ACE: adverse childhood experiences, Em: emotional, ROI: recurrence of illness, SB: suicidal behaviors, Pure BDI: sum of the cognitive Beck Depression Inventory items.

| Variables | PC06 | INT | PRO/ANTI | Total ACE |
| --- | --- | --- | --- | --- |
| INT | 0.583**<br>0.682** | - | - | - |
| PRO/ANTI | 0.351**<br>-0.007 | 0.243*<br>-0.094 | - | - |
| Total ACE | -0.021<br>0.352** | 0.138<br>0.284* | 0.287*<br>-0.078 | - |
| ACE EmAbuse | -0.085<br>0.458** | 0.142<br>0.265* | 0.321**<br>0.041 | - |
| ACE EmNeglect | 0.032<br>0.287* | 0.232<br>0.323** | 0.297*<br>0.049 | - |
| ROI | 0.043<br>0.614** | 0.244*<br>0.694** | 0.342**<br>-0.115 | 0.568**<br>0.352** |
| Neuroticism | 0.052<br>0.477** | 0.291*<br>0.602** | 0.307*<br>-0.128 | 0.583**<br>0.413** |
| LT_SB | 0.027<br>0.511** | 0.145<br>0.544** | 0.368**<br>-0.040 | 0.568**<br>0.275* |
| Current_SB | 0.073<br>0.297* | 0.034<br>0.374** | 0.321**<br>0.078 | 0.470**<br>0.134 |
| Pure BDI | -0.047<br>0.480** | 0.182<br>0.592** | 0.212<br>-0.147 | 0.580**<br>0.334** |
| Phenome | 0.023<br>0.539** | 0.298*<br>0.621** | 0.328**<br>-0.122 | 0.581**<br>0.421** |

### Prediction of clinical data using immune and atherogenicity data

Since we observed significant associations between INT and atherogenicity data, and since there are interactions between both kind of data, we constructed two new indices, namely a) the “atherommune” index which is a z composite score based on the sum of the z scores of both INT and PRO/ANTI indices; and b) the interaction pattern between INT x PRO/ANTI. Consequently, we introduced these two new variables, together with ApoE, RCT, BMI, ACE subtypes, and demographic data (age, sex, education) as input variables explaining clinical outcome data. **Table 4** shows that all clinical data were to a large extent predicted by the combined effects of these factors. Thus, ROI was associated with INT, INT x PRO/ANTI, three types of ACEs, and male sex. All these explanatory variables were positively associated with ROI and explained together 47.4% of its variance. We found that 37.7% of the variance in LT_SB (regression #2) was explained by the combined effects of atherommune and two ACE subtypes (all positively) and age and RCT (both inversely associated). Regression #3 displays that 29.4% of the variance in Current_SB is explained by two ACE subtypes, ApoE and INT x PRO/ANTI. A larger part of the variance in the pure BDI score (regression #4) was explained by three ACE subtypes, INT x PRO/ANTI (all positively associated) and age (inversely correlated). Up to 40.4% of the LT_phenome (regression # 5) was associated with emotional abuse, Atherommune, and INT x PRO/ANTI (all positively associated). We found that 46.2% in the current phenome (regression #6) was predicted by three ACE subtypes, atherommune, and INT x PRO/ANTI. **Figure 1** shows the partial regression of the current phenome of OMDD on the atherommune index. Regression #7 shows that a large part (58.3%) of the variance in the LT+Current phenome was associated with three ACE subtypes, INT, INT x PRO/ANTI, ApoE (all positively associated) and age (inversely associated).

**Figure 1.**
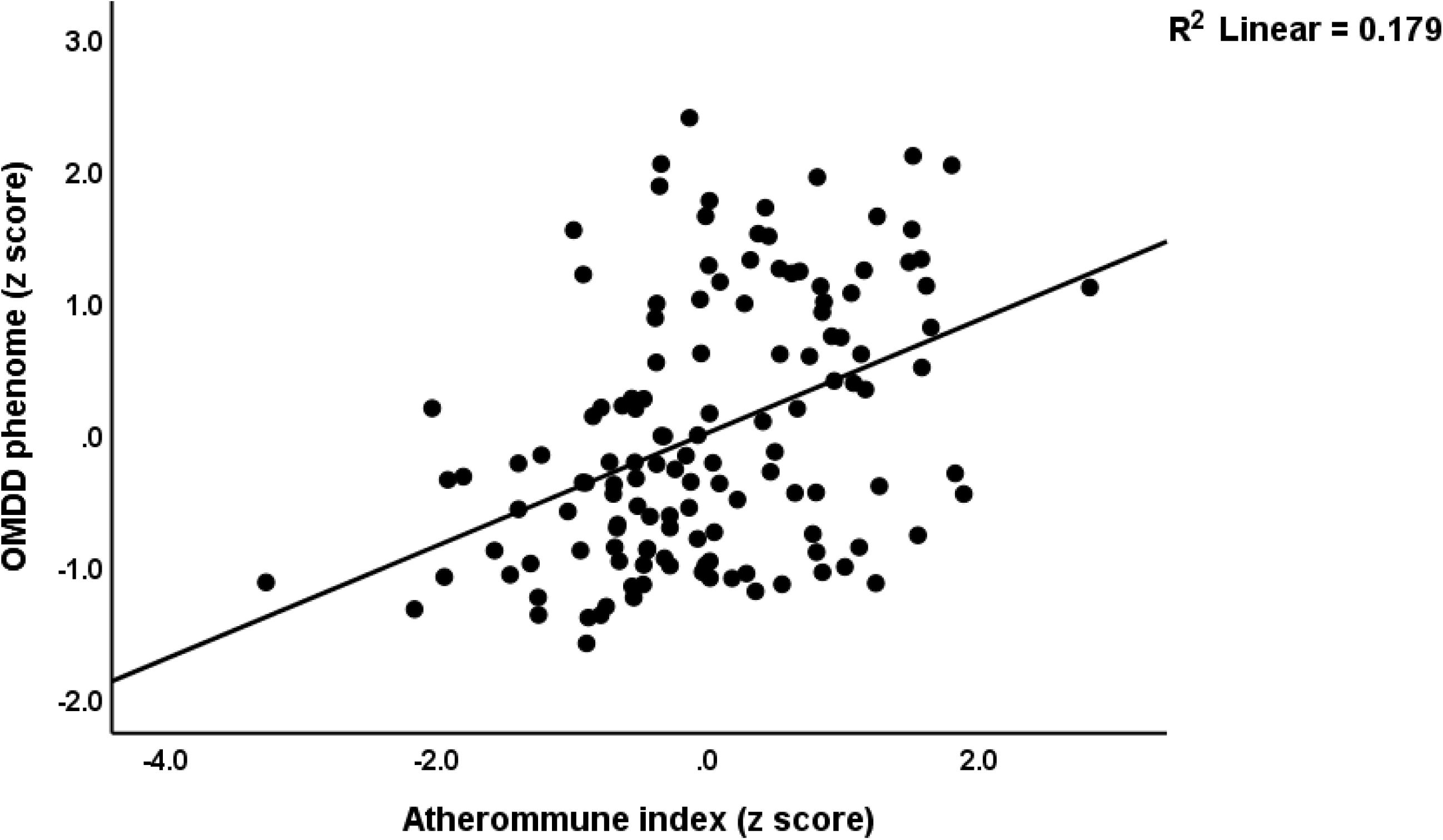
Partial regression of the current phenome of outpatient major depressive disorder on the atherommune index (p<0.001).

**Table 4.**
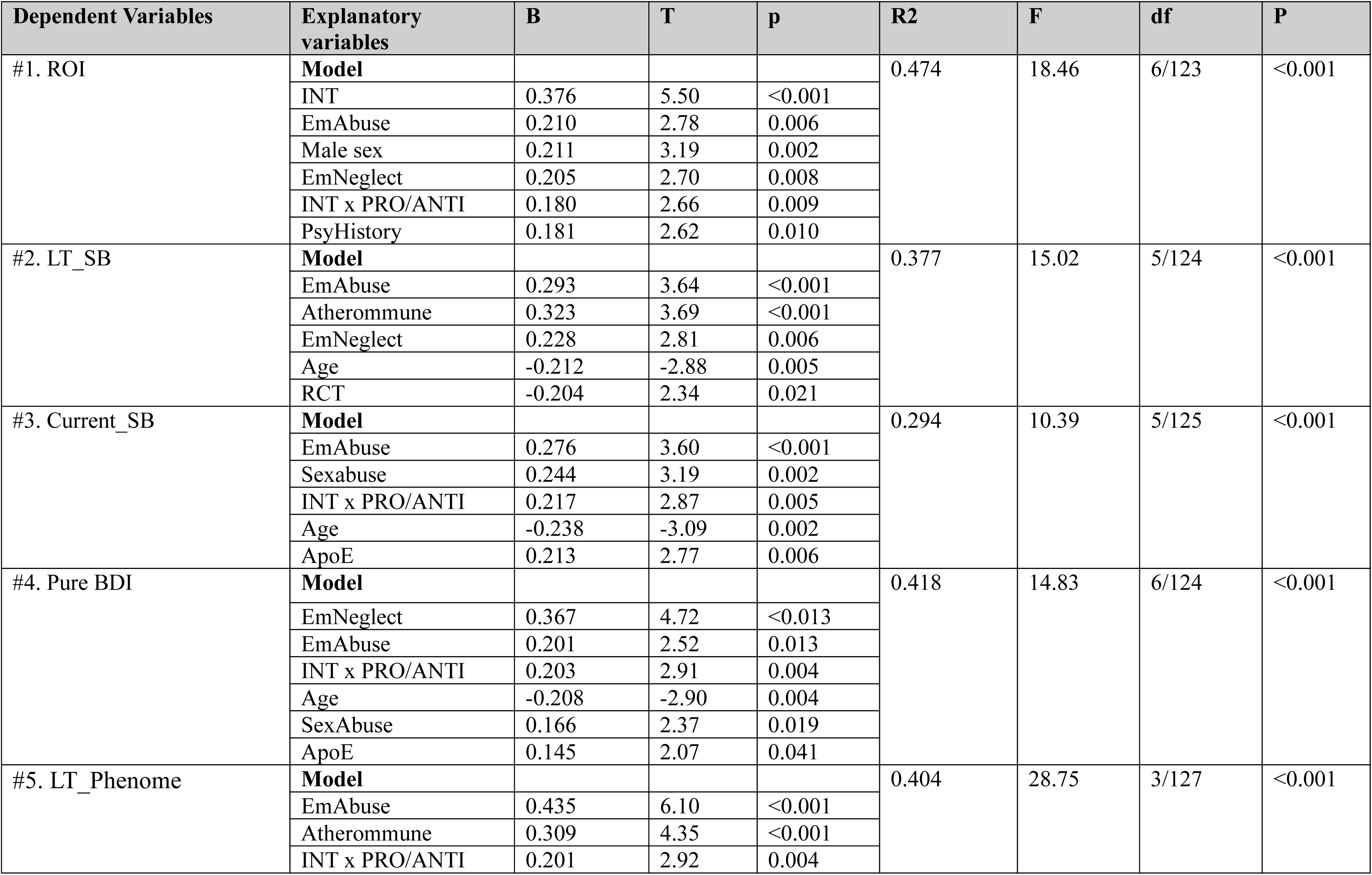
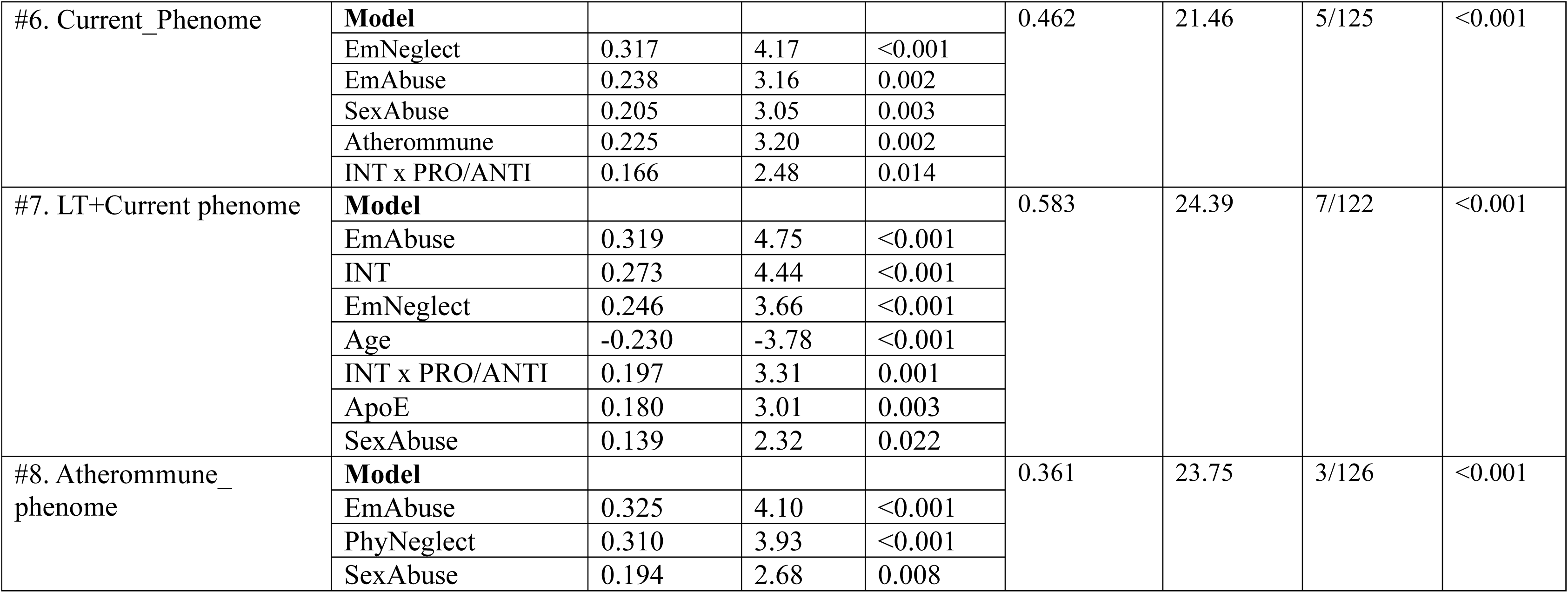
Results of multiple regression analysis with clinical phenome data as output variables and immune and atherogenicity data as input variables. ROI: recurrence of illness, INT: immune-linked neurotoxicity, ACE: adverse childhood experiences, Em: emotional, Phy: physical, Sex: sexual, Psy: psychiatric, PRO/ANTI: a composite score reflecting a comprehensive atherogenicity index, Atherommune: an index based on INT and PRO/ANTI, LT: lifetime, SB: suicidal behaviors, Pure BDI: sum of the cognitive Beck Depression Inventory items, Apo: apolipoprotein.

| Dependent Variables | Explanatory variables | B | T | p | R2 | F | df | P |
| --- | --- | --- | --- | --- | --- | --- | --- | --- |
| #1. ROI | **Model** |  |  |  | 0.474 | 18.46 | 6/123 | <0.001 |
|  | INT | 0.376 | 5.50 | <0.001 |  |  |  |  |
|  | EmAbuse | 0.210 | 2.78 | 0.006 |  |  |  |  |
|  | Male sex | 0.211 | 3.19 | 0.002 |  |  |  |  |
|  | EmNeglect | 0.205 | 2.70 | 0.008 |  |  |  |  |
|  | INT x PRO/ANTI | 0.180 | 2.66 | 0.009 |  |  |  |  |
|  | PsyHistory | 0.181 | 2.62 | 0.010 |  |  |  |  |
| #2. LT_SB | **Model** |  |  |  | 0.377 | 15.02 | 5/124 | <0.001 |
|  | EmAbuse | 0.293 | 3.64 | <0.001 |  |  |  |  |
|  | Atherommune | 0.323 | 3.69 | <0.001 |  |  |  |  |
|  | EmNeglect | 0.228 | 2.81 | 0.006 |  |  |  |  |
|  | Age | -0.212 | -2.88 | 0.005 |  |  |  |  |
|  | RCT | -0.204 | 2.34 | 0.021 |  |  |  |  |
| #3. Current_SB | **Model** |  |  |  | 0.294 | 10.39 | 5/125 | <0.001 |
|  | EmAbuse | 0.276 | 3.60 | <0.001 |  |  |  |  |
|  | Sexabuse | 0.244 | 3.19 | 0.002 |  |  |  |  |
|  | INT x PRO/ANTI | 0.217 | 2.87 | 0.005 |  |  |  |  |
|  | Age | -0.238 | -3.09 | 0.002 |  |  |  |  |
|  | ApoE | 0.213 | 2.77 | 0.006 |  |  |  |  |
| #4. Pure BDI | **Model** |  |  |  | 0.418 | 14.83 | 6/124 | <0.001 |
|  | EmNeglect | 0.367 | 4.72 | <0.013 |  |  |  |  |
|  | EmAbuse | 0.201 | 2.52 | 0.013 |  |  |  |  |
|  | INT x PRO/ANTI | 0.203 | 2.91 | 0.004 |  |  |  |  |
|  | Age | -0.208 | -2.90 | 0.004 |  |  |  |  |
|  | SexAbuse | 0.166 | 2.37 | 0.019 |  |  |  |  |
|  | ApoE | 0.145 | 2.07 | 0.041 |  |  |  |  |
| #5. LT_Phenome | **Model** |  |  |  | 0.404 | 28.75 | 3/127 | <0.001 |
|  | EmAbuse | 0.435 | 6.10 | <0.001 |  |  |  |  |
|  | Atherommune | 0.309 | 4.35 | <0.001 |  |  |  |  |
|  | INT x PRO/ANTI | 0.201 | 2.92 | 0.004 |  |  |  |  |
| #6. Current_Phenome | **Model** |  |  |  | 0.462 | 21.46 | 5/125 | <0.001 |
|  | EmNeglect | 0.317 | 4.17 | <0.001 |  |  |  |  |
|  | EmAbuse | 0.238 | 3.16 | 0.002 |  |  |  |  |
|  | SexAbuse | 0.205 | 3.05 | 0.003 |  |  |  |  |
|  | Atherommune | 0.225 | 3.20 | 0.002 |  |  |  |  |
|  | INT x PRO/ANTI | 0.166 | 2.48 | 0.014 |  |  |  |  |
| #7. LT+Current phenome | **Model** |  |  |  | 0.583 | 24.39 | 7/122 | <0.001 |
|  | EmAbuse | 0.319 | 4.75 | <0.001 |  |  |  |  |
|  | INT | 0.273 | 4.44 | <0.001 |  |  |  |  |
|  | EmNeglect | 0.246 | 3.66 | <0.001 |  |  |  |  |
|  | Age | -0.230 | -3.78 | <0.001 |  |  |  |  |
|  | INT x PRO/ANTI | 0.197 | 3.31 | 0.001 |  |  |  |  |
|  | ApoE | 0.180 | 3.01 | 0.003 |  |  |  |  |
|  | SexAbuse | 0.139 | 2.32 | 0.022 |  |  |  |  |
| #8. Atherommune_phenome | **Model** |  |  |  | 0.361 | 23.75 | 3/126 | <0.001 |
|  | EmAbuse | 0.325 | 4.10 | <0.001 |  |  |  |  |
|  | PhyNeglect | 0.310 | 3.93 | <0.001 |  |  |  |  |
|  | SexAbuse | 0.194 | 2.68 | 0.008 |  |  |  |  |

### Results of principal component analysis

Since a large part of the LT and Current phenome were associated with the Atherommune data, we have examined whether a validated factor could be extracted from LT_phenome, ROI, Current_phenome, INT, and Atherommune. **Table 5** shows that the principal component could be extracted from these five variables. The first component could be validated as a reliable latent construct with high VE (>50%), adequate Kaiser-Meyer-Olkin (KMO) test and a Cronbach’s alpha of 0.857. As such, the LT and Current phenome, ROI, and immune and atherogenicity pathways are manifestations of an underlying concept, labeled as the Atherommune-phenome. Table 4 shows that this principal component score was strongly predicted by three ACE subtypes which together explained 36.1% of the variance. **Figure 2** shows the partial regression of the atherommune-phenome of OMDD on mental neglect (p<0.001).

**Table 5.** Results of principal component (PC) analysis Lifetime phenome: first PC extracted from neuroticism, lifetime anxiety disorders and dysthymia, ROI: recurrence of illness, INT: immune-linked neurotoxicity, atherommune: composite score based on INT and a comprehensive atherogenicity index. KMO: Kaiser-Meyer-Olkin test.

| Indicators | PC loadings |
| --- | --- |
| Lifetime phenome | **0.803** |
| ROI | **0.835** |
| Current Phenome | **0.851** |
| INT | **0.754** |
| Atherommune | **0.741** |
| Variance explained | 63.68% |
| KMO | 0.738 |
| Bartlett's Test of Sphericity | 374.794 (df=10), $p<0.001$ |
| Cronbach's alpha | 0.857 |

**Figure 2.**
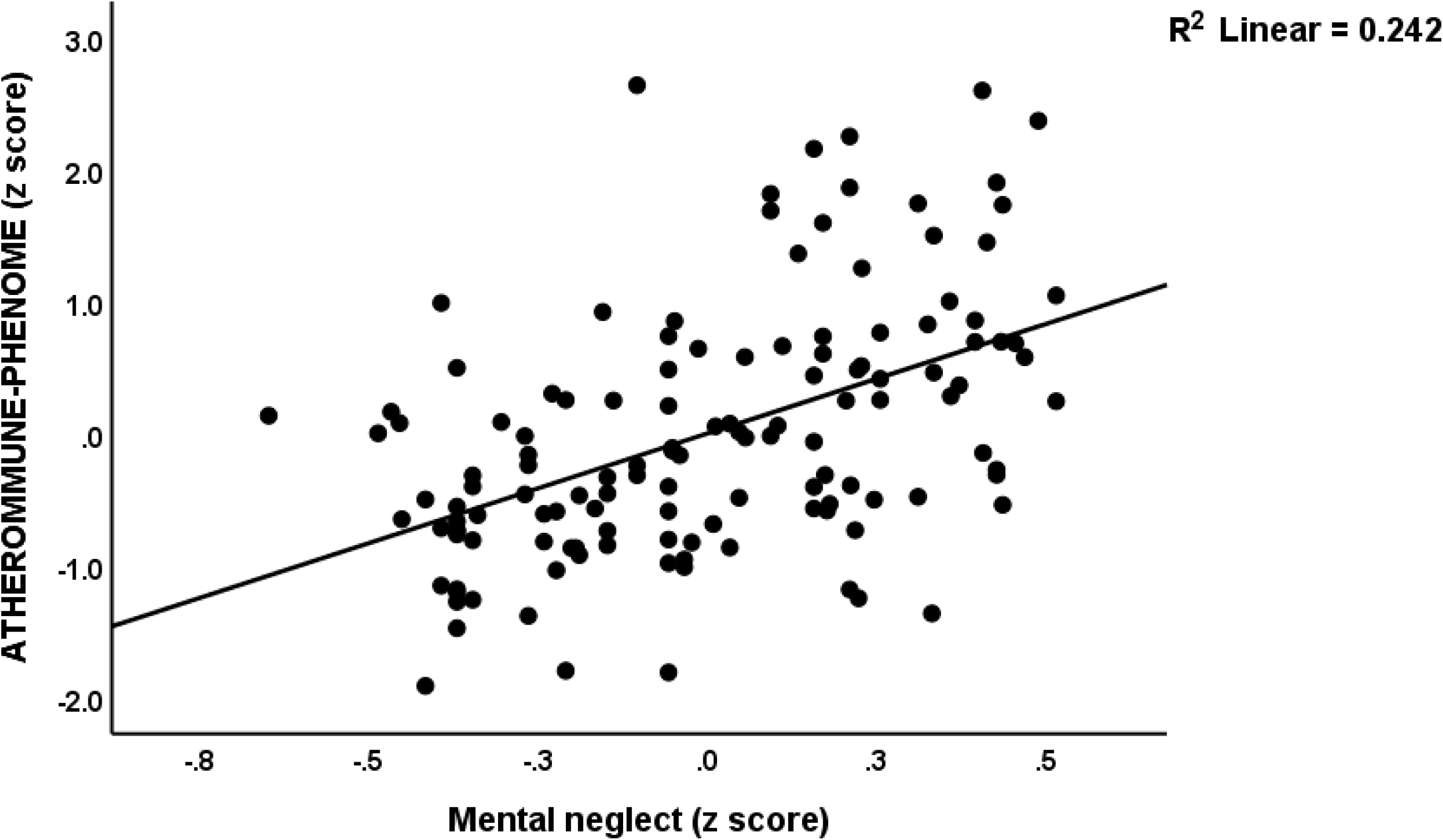
Partial regression of the atherommune-phenome of outpatient major depressive disorder on mental neglect (p<0.001).

## Discussion

### Associations between lipid metabolism and immune networks

The first main finding of this study is that, in subjects without MetS, the upregulated immune networks observed in OMDD are substantially associated with lipid metabolism biomarkers, including the free cholesterol/RCT ratio, the PRO/ANTI ratio, ApoE, and ApoB. Consequently, aberrations in lipid metabolism exhibit a robust correlation with the activation of immune networks. Conversely, the same immune networks were found to be substantially predicted by ACEs, such as sexual abuse and emotional neglect, in individuals with MetS. These findings indicate that the immune networks are influenced by different forces contingent upon the presence of MetS: lipid metabolism in the absence of MetS and ACEs in the presence of MetS. Nevertheless, these distinct impacts are remarkably simple to comprehend. Metabolic disorders may be so prevalent during MetS that the immune-lipid interactions (which may be observed in those without MetS) are not discernable in this restricted study sample.

Previously it was found that even very mild FE-OMDD in young individuals without MetS and any indicants of subclinical MetS is accompanied by elevated levels of free cholesterol and reduced levels of LCAT [24]. This suggests that FE-OMDD is a pre-proatherogenic condition. It can be inferred that as the disease progresses from FE-OMDD to more frequent episodes, there is an increase in pro-atherogenic indices (ApoE, ApoB, and free cholesterol/RCT) in OMDD in the absence of MetS. This elevation in these indices ultimately impacts immune functions and leads to heightened INT. In this condition, the direct effects of ACEs may not be detected as their effects on the phenome are mediated by the developing lipid aberrations [14].

Several aspects of lipid metabolism, including elevated cholesterol and ApoE levels, modulate the immune system, as described in the introduction [32, 28, 29, 33, 30, 31]. Furthermore, ApoE exerts an influence on oxidative, inflammatory, and immune responses, which extends to neuroimmune disorders like multiple sclerosis [44, 45]. CCL5 and CCL11 may be regulated by LDL-cholesterol [30, 46]. These findings support the current conclusion that lipid metabolism is associated with immune network functions. Indeed, an essential element in facilitating a proper immune response is cholesterol homeostasis, a process known as “cholesterol immunometabolism” [30]. The mechanism by which activated immune cells undergo metabolic reprogramming is facilitated by the upregulation of cholesterol and fatty acid synthesis, which converts the cells’ energy source to aerobic glycolysis [30]. An inflammatory immune cell phenotype is induced by elevated cholesterol levels in the plasma membrane[30]. Low grade inflammatory conditions (e.g., obesity and type 2 diabetes), as well as severe inflammatory conditions (e.g., rheumatoid arthritis and sepsis), are both characterized by intersections of lipid and immune responses. Hence, it is unsurprising that deviations in cholesterol immunometabolism are prevalent characteristics of the most severe human disorders in developed nations [47, 30], including OMDD, as demonstrated in the present study. Furthermore, the elevated atherogenicity indices identified in our research indicate that inflammatory processes might have been instigated by lipid reprogramming in macrophages [32, 48]. In addition, elevated oxysterol concentrations influence Th cells and facilitate the differentiation of Th17 cells and the trafficking of Th cells into the central nervous system (CNS) in animal models of multiple sclerosis[49, 50].

In addition to the immune network-affecting effects of free cholesterol, RCT, ApoB, and ApoE, we identified three distinct types of ACEs that have a profound effect on the immune networks. In a prior study [16] demonstrated that ACEs influence immune networks and are linked to classical M1 macrophage, Th1, Th1 polarization, IRS, and neurotoxicity immune profiles in FE-IMDD. However, these authors found no association between ACEs and the alternative M2 and Th2 immune profiles. In addition [16] identified strong correlations between ACEs and distinct cytokines, chemokines, and growth factors, particularly PDGF, IL-16, CCL27, and stem cell growth factor. Indeed, a complex network of cytokines, chemokines, and growth factors partially mediated the effects of ACEs on the phenome of depression [16]. A study conducted by [21] identified an association between ACEs and elevated production of M1, Th1, Th2, Th17, IRS, and INT profiles in diluted whole blood culture supernatant stimulated with PHA+LPS in a sample of individuals with extremely severe MDD. Those measurements are indicative of immune profile sensitization and the responsiveness of the immune system subsequent to injuries [51]. This suggests that ACEs induce immune sensitization in a tight network of cytokines, chemokines, and growth factors [21]. Moreover, the latter network’s ACE-associated sensitization is mediated by JAK-STAT, nuclear factor-B, G-protein coupled receptor, PI3K/Akt/RAS/MAPK, and hypoxia signaling, according to enrichment analysis [21]. Additionally, prior investigations have demonstrated that ACEs have the potential to impact acute phase proteins such as C-reactive protein (CRP) [52].

In conclusion, in OMDD, it appears that increased levels of free cholesterol, ApoB and ApoE, and ACEs modulate immune networks, resulting in increased INT.

### Neurotoxicity through lipid metabolism and immune activation in OMDD

The clinical phenome features of OMDD (ROI, including lifetime suicidal behaviors), the lifetime phenome (neuroticism + lifetime anxiety disorders + dysthymia), and the current phenome (including current suicidal behaviors) were found to be strongly correlated with immune networks and INT only in individuals without MetS. In contrast, only a limited number of associations with a smaller effect size were observed in individuals with MetS. Conversely, individuals lacking MetS exhibited significant impacts of atherogenicity indices on the features of the OMDD phenome. Nevertheless, the phenome characteristics of individuals with MetS and those without MetS were impacted by ACEs. As previously mentioned, in order to assess the impact of atherogenicity on immune function and the phenome, it is necessary to examine patients without MetS. This is due to the fact that the study sample containing MetS is inherently a restricted sample, consisting solely of individuals with metabolic disorders. Furthermore, a previous study demonstrated that interactions between MetS and particular biomarkers (e.g., CCL5, VEGF) can predict the phenotypic characteristics of OMDD [25]. Consequently, the disparities between individuals with and without MetS regarding the associations between lipid metabolism, INT, and phenotypic characteristics are expected.

It is crucial to underscore that the elevated atherogenicity and reduced RCT indices employed in this investigation may not only result in increased atherogenicity but also increased neurotoxic effects. First, an excess of cholesterol may impair the blood-brain barrier [53, 54] and, as mentioned earlier, stimulate the immune system, resulting in increased transport and activation of Th cells into the CNS. An additional characteristic of neurons with excessive cholesterol accumulation is heightened susceptibility to NMDA excitotoxicity [55]. Furthermore, an overabundance of cholesterol accumulation and the subsequent production of oxysterols may trigger neuroinflammation, heightened oxidative stress, and impaired mitochondrial functions - all of which are significant contributors to neurodegenerative disorders. Overabundance of triglycerides, cholesterol, and ApoB may be exacerbated by elevations in ApoE [24]. Reduced LCAT activity may result in diminished RCT, which in turn may lead to elevated levels of free cholesterol and accelerated atherogenicity progression [24]. LCAT is an essential factor in the maturation of HDL-cholesterol and exhibits antioxidant properties [14]. Consequently, decreased LCAT activity could potentially contribute to the heightened oxidative stress accompanied by increased aldehyde production in MDD, thereby potentially exacerbating neurotoxicity [24].

In summary, the immune networks and lipid profiles (atherogenicity and decreased RCT and LCAT) that have been identified in OMDD may each contribute to enhanced neurotoxicity via unique mechanisms. Consequently, we devised two novel indices that consider the interdependent effects of INT and atherogenicity, namely the “atherommune index,” which is a composite of the two factors, or the interaction term between the two.

### ACEs, atherogenicity, immune-linked neurotoxicity and the OMDD phenome

The third significant discovery of this research is that a combination of atherogenic and INT biomarkers in conjunction with ACEs, specifically emotional neglect and abuse, and sexual abuse, could predict the majority of the features of OMDD. In the past, it was documented that the impact of ACEs on the phenome was partially mediated by increased INT, decreased CIRS activities (particularly IL-4 levels), and atherogenicity [21, 16, 14]. Importantly, atherogenicity, neurotoxicity, and the interaction of atherogenicity and INT all predicted the phenotypic characteristics of OMDD, including ROI, lifetime and current suicidal behaviors, and lifetime and current phenotype of OMDD, in this study. Most importantly, it was discovered that a single principal component could be derived from ROI (including number of episodes, and lifetime suicidal behaviors), the lifetime phenome (including neuroticism, dysthymia, and any anxiety disorders), the current phenome (including severity of depression, anxiety and current suicidal behaviors), immune-mediated neurotoxicity, and the atherommune index. This indicates that the illness OMDD is the underlying latent construct through which all features of MDD manifest, including INT and its interactions with atherogenicity. In order to model OMDD, we have therefore developed a novel pathway-phenotype and labeled it as the "atherommune-phenome" pathway-phenotype. Consequently, activated INT and the intersection of the latter with increasing atherogenicity comprises the essence of OMDD and all of its features. Further, the phenotype of this pathway is significantly predicted by three distinct adverse childhood experiences (ACEs): emotional abuse and neglect, sexual abuse, and neglect.

### Limitations

The investigation would have been further enhanced had oxysterols been quantified as well as other biomarkers of oxidative stress. Further investigation is warranted to explore the correlations between lipid metabolism and INT across the various phenotypes of MDD, such as FE-OMDD, FE-IMDD, and multiple episode IMDD.

## Conclusions

A significant portion (58.3%) of the variance in the lifetime + current phenome features of OMDD was accounted for by INT, interactions between the latter and atherogenicity, ApoE, three ACE subtypes (all positively), and age (inversely). A single validated latent construct could be derived from several factors such as ROI, the lifetime phenome, the current phenome, immune-liked neurotoxicity, and atherommune index. Three ACE subtypes accounted for 36.1% of the variance in this factor. As such, we have developed a new pathway phenotype called the "atherommune-phenome" to represent OMDD. This phenotype highlights the importance of INT and its interactions with atherogenicity in OMDD.

The findings indicate that both activated immune networks with increased INT and the interactions between the latter and lipid profiles are potential targets for new drugs treatments in OMDD. Thus, not only targeting the activated immune pathways but also reprogramming of lipid metabolism or blockade of cholesterol biosynthesis may be new drug targets to treat OMDD. There is existing evidence suggesting that statins may possess antidepressant properties, as indicated by studies conducted by [56–58]. However, it is recommended to complement such treatment with coenzyme Q10, as suggested by Maes et al. [59]. Further research is needed to investigate the impact of using statins and coenzyme Q10 together on INT, as well as the relationship between the latter and atherogenicity indices in MDD, and how this relates to its clinical effectiveness.

## Author**’**s contributions

Conceptualization and study design: MM and JK; first draft writing: MM; statistical analysis: MM; editing: all authors; recruitment of patients: JK. All authors approved the last version of the manuscript.

## Ethics approval and consent to participate

The research project (#445/63) was approved by the Institutional Review Board of Chulalongkorn University’s institutional ethics board. All patients and controls gave written informed consent prior to participation in the study.

## Funding

This work was supported by the Ratchadapiseksompotch Fund, Graduate Affairs, Faculty of Medicine, Chulalongkorn University (Grant number GA64/21), a grant from CU Graduate School Thesis Grant, and Chulalongkorn University Graduate Scholarship to Commemorate the 72nd Anniversary of His Majesty King Bhumibol Adulyadej to KJ; the Thailand Science research, and Innovation Fund Chulalongkorn University (HEA663000016), and a Sompoch Endowment Fund (Faculty of Medicine) MDCU (RA66/016) to MM, and Grant № BG-RRP-2.004-0007-С01 „Strategic Research and Innovation Program for the Development of MU - PLOVDIV–(SRIPD-MUP)“, Creation of a network of research higher schools, National plan for recovery and sustainability, European Union – NextGenerationEU.

## Conflict of interest

The authors have no commercial or other competing interests concerning the submitted paper.

## Data Availability Statement

The dataset generated during and/or analyzed during the current study will be available from the corresponding author upon reasonable request and once the dataset has been fully exploited by the authors.

## Notes

### Competing Interest Statement

The authors have declared no competing interest.

