## supplementary file for "Are abnormalities in lipid metabolism, together with adverse childhood experiences, the silent causes of immune-linked neurotoxicity in major depression?"

**ESF, Table 1**. Overview of the cytokines, chemokines, and growth factors measured in the current study

| Protein abbreviations | Gene Symbol | > OOR (%) | Protein name / alias |
|---|---|---|---|
| **IFN-α2** | **IFNA2** | 60.8 | Interferon-α2 |
| **IFN-γ** | **IFNG** | 81.9 | Interferon-γ |
| **IL-1α** | **IL1A** | 60.8 | Interleukin-1α |
| **IL-1β** | **IL1B** | 78.3 | Interleukin-1β |
| **sIL-1RA** | **IL1RN** | 100 | Soluble interleukin-1 receptor antagonist |
| **IL-2R** | **IL2RA** | 100 | Soluble interleukin-2 receptor |
| **IL-4** | **IL4** | 100 | Interleukin-4 |
| **IL-9** | **IL9** | 100 | Interleukin-9 |
| **IL-10** | **IL10** | 56.4 | Interleukin-10 |
| **IL-12p40** | **IL12B** | 60.8 | Interleukin-12 p40 |

| | | | |
|---|---|---|---|
| **IL-13** | **IL13** | 68.1 | Interleukin-13 |
| **IL-15** | **IL15** | 10.9 | Interleukin-15 |
| **IL-16** | **IL16** | 100 | Interleukin-16 |
| **IL-17** | **IL17A** | 60.8 | Interleukin-17 |
| **IL-18** | **IL18** | 100 | Interleukin-18 |
| **TNF-α** | **TNF** | 100 | Tumor necrosis factor-α |
| **TNF-β** | **LTA** | 100 | Tumor necrosis factor-β or lymphotoxin-alpha (LT-α) |
| **TRAIL** | **TNFSF10** | 100 | TNF-related apoptosis-inducing ligand (TRAIL) or tumor necrosis factor ligand superfamily member 10 (TNFSF10) |
| **LIF** | **LIF** | 94.2 | Leukemia inhibitory factor |
| **MIF** | **MIF** | 100 | Macrophage migration inhibitory factor-like protein (MIF) or glycosylation-inhibiting factor |
| **G-CSF** | **CSF3** | 100 | Granulocyte colony stimulating factor (G-CSF) or colony stimulating factor 3 (CSF3) |
| **M-CSF** | **CSF1** | 100 | Macrophage colony-stimulating factor (M-CSF) or colony stimulating factor 1 (CSF1) |
| **GM-CSF** | **CSF2** | 60.8 | Granulocyte-macrophage colony-stimulating factor (GM-CSF) or colony-stimulating factor 2 (CSF2) |
| **CCL2 or MCP1** | **CCL2** | 100 | C-C motif chemokine ligand 2 (CCL2) or monocyte chemoattractant protein 1 (MCP1) |
| **CCL3 or MIP-1α** | **CCL3** | 100 | C-C motif Chemokine ligand 3 (CCL3) or macrophage inflammatory protein 1-alpha (MIP-1α) |

| | | | |
|---|---|---|---|
| **CCL4 or MIP-1β** | **CCL4** | 100 | C-C motif chemokine ligand 4 (CCL4) or macrophage inflammatory protein 1β (MIP-1β) or lymphocyte activation gene 1 protein |
| **CCL5 or RANTES** | **CCL5** | 100 | C-C motif chemokine ligand 5 (CCL5) or regulated upon activation, normally T-expressed, and presumably Secreted (RANTES) |
| **CCL7 or MCP3** | **CCL7** | 60.8 | C-C motif chemokine ligand 7 (CCL7) or monocyte-chemotactic protein 3 (MCP3). |
| **CCL11 or Eotaxin** | **CCL11** | 100 | C-C motif chemokine ligand 11 (CCL11) or eosinophil chemotactic protein |
| **CCL27 or CTACK** | **CCL27** | 100 | C-C motif chemokine ligand 27 (CCL27) or cutaneous T-cell attracting chemokine (CTACK) |
| **CXCL1 or GRO-α** | **CXCL1** | 100 | C-X-C motif chemokine 1 (CXCL1) or growth-regulated alpha protein (GRO) |
| **CXCL8 or IL-8** | **CXCL8** | 100 | C-X-C motif chemokine ligand 8 (CXCL8) or interleukin-8 (IL-8) |
| **CXCL9 or MIG** | **CXCL9** | 100 | C-X-C motif chemokine ligand 9 (CXCL9) or monokine induced by gamma interferon (MIG) |
| **CXCL10 or IP10** | **CXCL10** | 100 | C-X-C motif chemokine ligand 10 (CXCL10) or Interferon gamma-induced protein 10 (IP10) |
| **CXCL12 or SDF-1α** | **CXCL12** | 100 | C-X-C motif chemokine 12 (CXCL12) or stromal cell-derived factor 1 (SDF-1α) |
| **FGF** | **FGF2** | 91.3 | Fibroblast growth factor 2 (FGF) or basic fibroblast growth factor |
| **HGF or SF** | **HGF** | 100 | Hepatocyte growth factor (HGF) or scatter factor (SF) |
| **BNGF** | **NGF** | 63.8 | β-nerve growth factor (NGF) |
| **PDGF** | **PDGFA** | 100 | Platelet derived growth factor (PDGF) |
| **SCF** | **KITLG** | 100 | Stem cell factor (SCF) or Kit ligand (KITLG) |

| | | | |
|---|---|---|---|
| **SCGF-β or CLEC11A** | **CLEC11A** | 100 | Stem cell growth factor (SCGF) or C-type lectin domain family 11 member A (CLEC11A) |
| **VEGF** | **VEGFA** | 89.1 | Vascular endothelial growth factor (VEGF) |

Adapted from: *Maes M, Rachayon M, Jirakran K, Sodsai P, Klinchanhom S, Gałecki P, Sughondhabirom A, Basta-Kaim A. The Immune Profile of Major Dysmood Disorder: Proof of Concept and Mechanism Using the Precision Nomothetic Psychiatry Approach. Cells. 2022 Mar 31;11(7):1183. doi: 10.3390/cells11071183. PMID: 35406747; PMCID: PMC8997660.*

*Kalayasiri R, Dadwat K, Supaksorn T, ,Sirivichayakul S, Maes M. Methamphetamine (MA) use, MA dependence, and MA-induced psychosis are associated with increasing aberrations in the compensatory immunoregulatory system and interleukin-1α and CCL5 levels. medRxiv 2023.03.26.23287766; doi: https://doi.org/10.1101/2023.03.26.23287766*

### ESF, Table 2. Description of the immune profiles used in this study

| Immune Profile | Members |
|---|---|
| **PC05** | G-CSF, CXCL1, HGF, sIL-1RA, sIL-2R, IL-4, IL-8, IL-9, IL-16, IL-17, CCL2, MIF, CCL3, CCL4, PDGF, CCL5, SCGF, CXCL12, TNF-α, TNF-β |
| **PC06** | sIL-1RA, IL-4, IL-9, IL-17, CCL4, PDGF, CCL5, CXCL12, TNF-α, TNF-β |
| **PCdown** | CCL3, CSF1, VEGF, NGF, IL-10 |

PC: principal component

For construction of the PCs:

Maes M, Jirakran K, Vasupanrajit A, Zhou B, Tunvirachaisakul C, Almulla AF. Major depressive disorder, neuroticism, suicidal behaviors, and depression severity are all associated with neurotoxic immune networks and their intricate interactions with metabolic syndrome. medRxiv 2024.01.20.24301553; doi: https://doi.org/10.1101/2024.01.20.24301553
